# GASTROINTESTINAL MUCOSAL DAMAGE AND SUBSEQUENT RISK OF PARKINSON’S DISEASE

**DOI:** 10.1101/2024.02.29.24303572

**Authors:** Jocelyn J. Chang, Subhash Kulkarni, Trisha S. Pasricha

## Abstract

**Introduction:** The gut-first hypothesis of Parkinson’s Disease (PD) has gained traction, yet the inciting events triggering PD from gut-related factors remain unclear. While H. pylori infection is linked to peptic injury and is 1.47 times more prevalent in PD individuals, it is unknown how gastrointestinal mucosal damage (MD) may increase the risk of PD. We aimed to study the association between upper endoscopy findings of MD and subsequent PD development.

**Methods:** In our retrospective study of 18,305 adults without prior PD, undergoing upper endoscopy between 2000 and 2005, patients with MD were matched with non-MDs. PD risk in MDs versus non-MDs was assessed using incidence rate ratio (IRR) and multivariate Cox analysis, controlling for covariates.

**Results:** In the matched cohort, MD patients were significantly more likely to develop PD (IRR 3.00, p<0.0001), even after covariate adjustment (HR 2.42, p<0.001). Covariates including constipation, dysphagia, older age, and male sex were also associated with higher PD risk. Among MDs, H. pylori presence (AOR 5.38, p=0.04) and chronic NSAID use (AOR 3.28, p=0.04) increased PD odds, while chronic smoking decreased PD odds (AOR 0.19, p<0.05).

**Conclusion:** MD elevates PD risk, with H. pylori increasing risk only in the presence of MD, suggesting a closer link between PD and gastric mucosa disruption. Furthermore, chronic NSAID use significantly raises PD odds in MD, while chronic smoking reduces PD risk in this context. Increased vigilance among MD patients for future PD risk is warranted, with further studies needed to elucidate precise pathophysiology.

## Introduction

Parkinson’s Disease (PD), a progressive neurodegenerative disorder, affects 8.5 million people worldwide based on 2019 estimates^1^, with prevalence more than doubling over the past 3 decades. Characterized by the degeneration of dopaminergic neurons in the substantia nigra, PD manifests through motor symptoms such as tremors, rigidity, and bradykinesia, as well as non-motor symptoms including constipation.

Braak’s hypothesis, rooted in neuropathological staging of PD, outlines a consistent progression pattern from specific induction sites^2^. Additionally, accumulating evidence indicates a prolonged prodromal phase of PD, lasting years and characterized by significant gastrointestinal (GI) dysfunction^3^. The convergence of these factors suggests a gut-first hypothesis, proposing that pathology originates in the gut and travels to the brain through the vagus nerve. Recent animal studies reinforce this concept, showing the transmission of gut-initiated pathology via the vagus nerve, leading to central nervous system neurodegeneration and motor deficits^4,5^. These findings provide substantial support for PD originating in the gut. However, the precise inciting factor triggering PD pathology in the gut remains elusive, complicating efforts to fully understand PD’s onset. A recent nationwide study found no association between a history of inflammatory bowel disease and subsequent risk of PD, suggesting that MD in the distal GI tract where pelvic rather than vagal innervation is the hallmark, may play a less important role in pathogenesis^6^.

In this context, Helicobacter pylori (*H. pylori)* infection, a well-known risk factor for upper GI tract ulcers and malignancies, has emerged as a point of interest. Epidemiological studies have reported a higher prevalence of *H. pylori* infection in PD patients compared to the general population, implying a possible association between *H. pylori* and PD^7-9^. The pathogen’s role in inducing GI mucosal damage (MD) and its systemic inflammatory responses present a compelling area for investigation in relation to PD.

Indeed, a significant gap remains in understanding the relationship between broader upper GI MD, such as peptic ulcer disease and erosions, and the development of PD as most studies to date have focused on correlational links to a history of *H pylori* infection. Additionally, while non-steroidal anti-inflammatory drug (NSAID) use is a known risk factor for peptic ulcer disease (PUD), existing literature is mixed regarding its role in PD. Some studies have suggested a possible neuroprotective effect of NSAIDs^10^ and an associated reduced risk of PD, while other studies^11,12^ found no evidence for association. We hypothesized that significant pathological defects to GI mucosa, including but not limited to those associated with *H. pylori* and NSAID use, may serve as an inciting event that could precipitate subsequent development of PD—though the net effect of NSAIDs on PD risk might be offset by its proposed neuroprotective properties.

Our study explores this gap by examining the association between GI MD, confirmed through upper endoscopy (EGD) findings, and the subsequent development of PD. By retrospectively analyzing a cohort of patients who underwent EGD, this study aims to develop a better mechanistic understanding of the potential gut-first pathway of PD pathogenesis, thereby contributing to a more comprehensive understanding of the disease and opening new avenues for early intervention and treatment strategies.

## Methods

### Retrospective cohort study design

We performed a retrospective cohort study to determine the risk of developing Parkinson’s disease among patients with a history of MD compared to those without. We then performed a nested case-control study to assess for specific covariates that might account for the associations observed in the broader cohort.

#### Patient selection

Patients were selected using Research Patient Data Registry (RPDR), a Partners Healthcare electronic database that stores data on patient demographics, encounters, medications, procedures, and billing codes. Using the RPDR search query tool, we identified a cohort of 18,305 adults based on the following inclusion criteria: (1) within the MGB system, (2) adults 18 years and older, (3) history of EGD with biopsy between 2000 and 2005, and (4) no history of PD (ICD10-G20.C) prior to initial EGD. Patients were excluded if they had missing data for a covariate of interest.

Patients with positive endoscopic findings for MD were matched with non-MD patients in a 1:3 ratio based on age, sex, and date of EGD. MD was defined as the presence of “erosion”, “esophagitis”, “ulcer”, or “peptic injury” observed on EGD or pathology reports. Final cohort size after matching included is shown in **Figure 1**.

**Figure 1:**
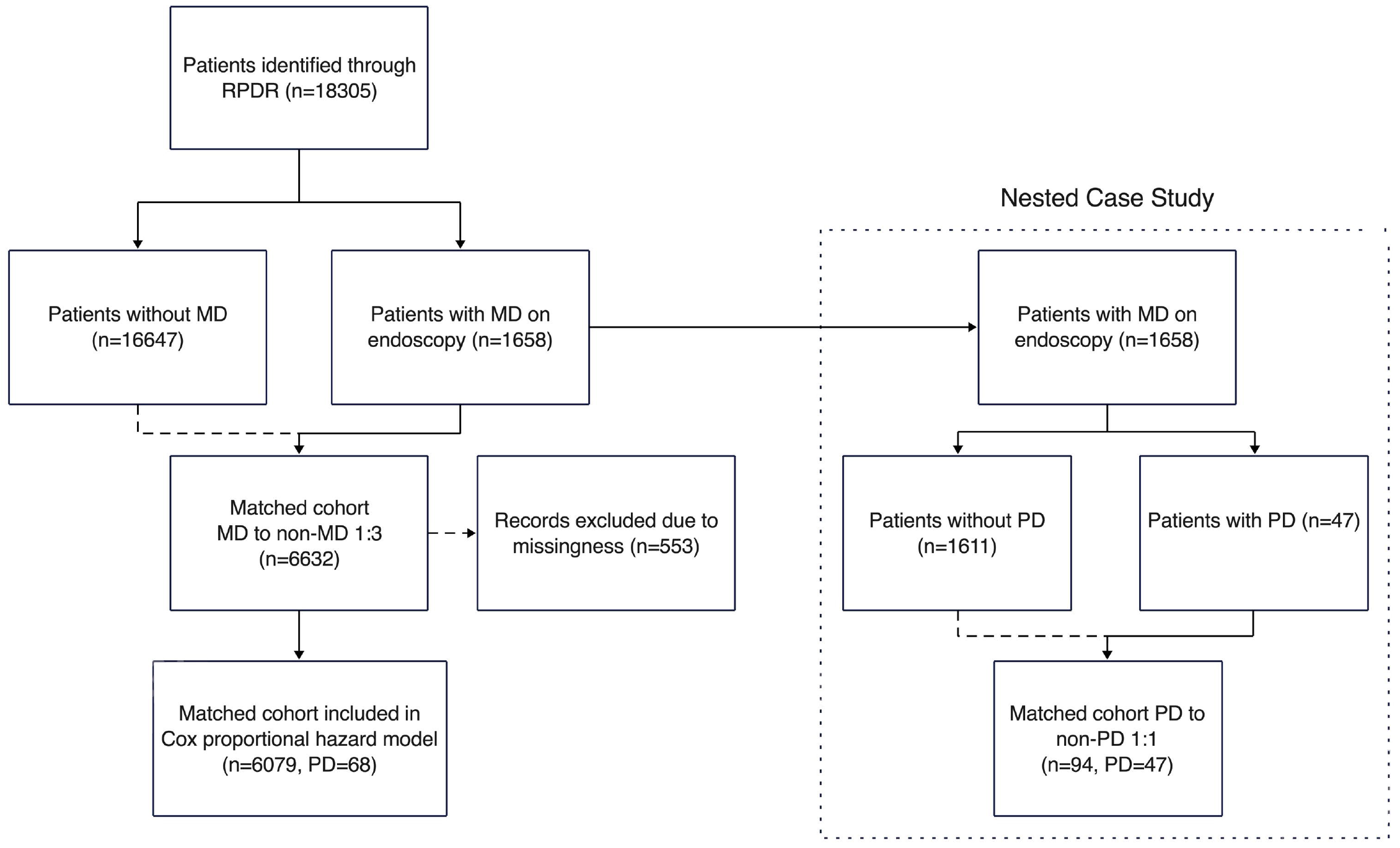
Patient Selection Process RPDR: Research Patient Data Registry; MD: mucosal damange; PD: Parkinson’s disease

#### Follow-up

Each participant was followed until (1) diagnosis of PD; (2) in the absence of a PD diagnosis, censoring at an outpatient appointment with no subsequent follow-up for greater than 2 years, representing loss to follow-up; or (3) death.

#### Outcomes and exposures

Cases of PD were identified through a ICD9:332.0 and ICD10:G20 code screen that was confirmed per medical record review.

To identify comorbidities, we used the enhanced version of the Charlson-Deyo comorbidity index (CCI) for administrative data. Other covariates were chosen based on their known or suspected links to peptic ulcer disease or PD, as suggested by existing literature^3,13,14^.

Age, sex, race, and BMI were extrapolated from demographic data. With regards to medications: long-term use of NSAIDs was identified using ICD9:V58.64 and ICD10:Z79.1; PPI use, dichotomized as any history or none, was extracted from medication prescription history. For substance use history: alcohol use disorder was defined by ICD9 codes 303- and 305.0-, and ICD10 equivalents F10-; history of complications related to alcohol use, including alcoholic gastritis and alcoholic liver disease, were defined by ICD9 codes 202, 535.3, 525.3, 571-, and 980, along with ICD10 equivalents K29.2, K70-; and chronic tobacco smoking history or exposure was identified through ICD9 codes 305.1- and E869.4, and ICD10 equivalents Z72.0, Z87.891, Z57.31, and Z77.22. For GI covariates: history of constipation, dysphagia, and GERD were defined per ICD9 codes 564.0-, 787.2-, 530.11 and 530.81 respectively, in addition to ICD10 equivalents K59.0-, R13.1-, and K21-respectively; and H. pylori infection was defined as the presence of H. pylori identified on endoscopic pathology stains.

#### Statistical analysis

Based on patient outcomes and exposures, descriptive characteristics of our cohort were reported as means and standard deviations for continuous variables, and total number and percent of total number for categorical variables. Descriptive data was then analyzed using t-tests and chi-square tests, respectively, with p-value ≤ 0.05 as statistically significant.

Incidence rate of PD in our matched cohort was calculated per 10,000 person-years and compared with the incidence of PD in the general US population for generalizability. We then calculated the incidence rate ratio (IRR) of PD in MD versus non-MD patients.

Multivariate Cox proportional hazard analysis was conducted in a stepwise manner to incrementally assess the impact of each covariate on the risk of developing PD, ensuring a thorough examination of their individual and combined effects without overfitting our model. We then assessed for possible violation of the proportional hazard’s assumption with log-log plots and Schoenfeld residuals, confirming that the assumption in our Cox model was not violated.

### Nested Case-Control Study

Building on our cohort study, a nested case-control analysis was conducted within the group of patients previously found to have mucosal damage (MD), assessing for specific factors that might account for the associations observed in the broader cohort.

#### Patient selection and covariates

In our nested cohort, we defined cases as patients with MD, subsequently diagnosed with PD. Controls were defined as patients with MD who did not later develop PD according to follow-up. Cases were matched with controls in a 1:1 ratio based on age, sex, and month of the case’s diagnosis by incidence density sampling. Covariates were determined in the same manner as in the retrospective study.

#### Statistical analyses

Crude and adjusted odds ratios with 95% confidence intervals were calculated using conditional logistic regression to determine the association between PD and specific covariates: chronic smoking, esophagitis, H. pylori, chronic NSAID use, PPI use, and GERD.

## Results

Total cohort size was 6079 patients, mainly White (81.1%), male (56.6%), and over 65 years of age (61.9%). In our cohort, no significant demographic differences were noted between MD and non-MD patients as a result of matching. With regard to covariates, there was significantly higher prevalence in MD compared to non-MD participants of: PPI use (81.5% vs 49.6%, p<0.0001), chronic NSAID use (20.0% vs 6.5%, p<0.0001), GERD (89.0% vs 67.6%, p<0.0001), chronic smoking (16.5% vs 12.6%, p<0.004), constipation (49.0% vs 27.4%, p<0.0001), and dysphagia (49.8% vs 30.3%, p<0.0001); and no significant difference in *H. pylori* infection rates or CCI in MD compared to non-MD participants (**Table 1**).

**Table 1.**
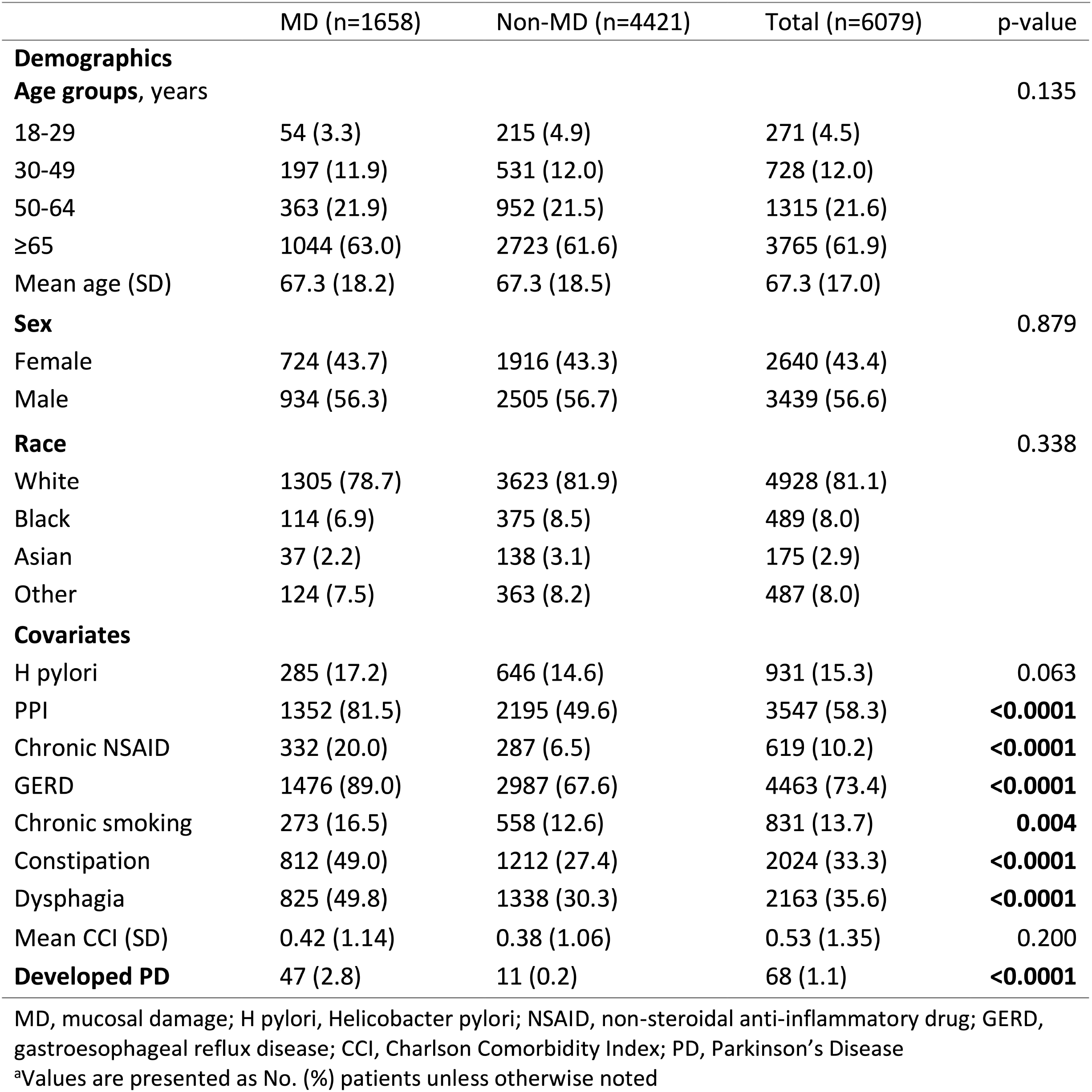
Patient Characteristics. matched 1:3 by age, sex, initial endoscopy date ^a^

Our results indicate a notably higher likelihood of PD development among patients with MD compared to those without MD (prevalence 2.8% vs 0.2%, p<0.0001; incidence rate ratio [IRR] 3.00 p<0.0001). After adjusting for covariates, the risk associated with MD remained pronounced (hazard ratio [HR] 2.42, 95% confidence interval [CI] 1.46-4.00, p<0.001). Of the 68 patients who later developed PD, 47 had MD on initial biopsy (69.1%). Overall incidence of PD in our cohort was 16 per 100,000 person-years, comparable with reported overall incidence of 17 per 100,000 person-years worldwide [hirsh, 2016]. Survival curves for time to PD diagnosis was less favorable for patients with MD than those without (**Figure 2**).

**Figure 2.**
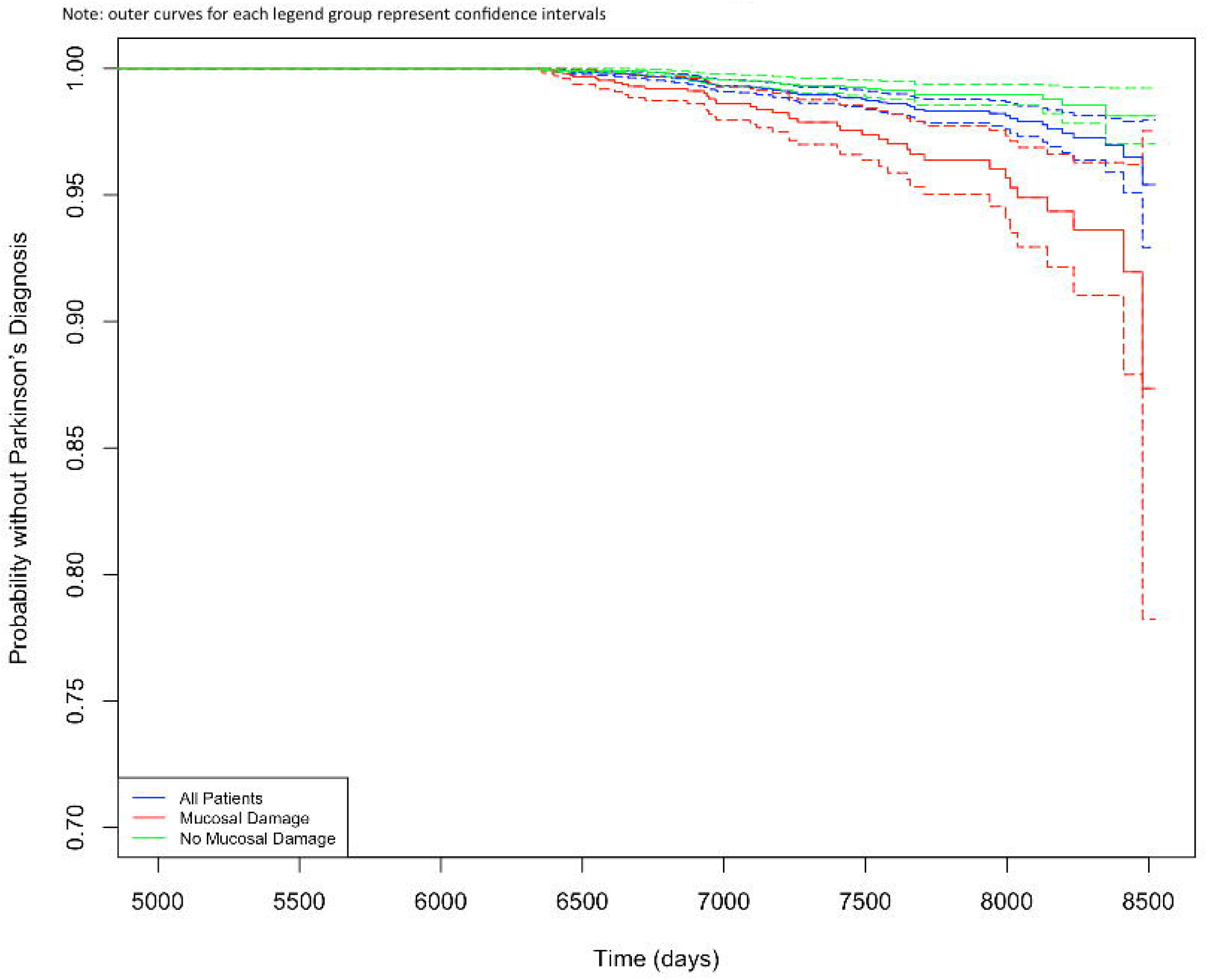
Survival Analysis:Time to Parkinson’s Diagnosis Note:outer curves for each legend group represent confidence intervals

Factors associated with an increased risk of developing PD included: age (1.03, 95% CI 1.01-1.05, p=0.003), sex (HR 0.60, 95% CI 0.37-0.98, p=0.04), constipation (HR 2.78, 95% CI 1.62– 4.76, p<0.001), and dysphagia (HR 2.45, 95% CI 1.44-4.16, p<0.001). Contrastingly, race, Charlson Comorbidity Index (CCI), and history of *H. pylori* infection on endoscopy were not associated with risk of PD (**Table 2a, Figure 3a**).

**Table 2.**
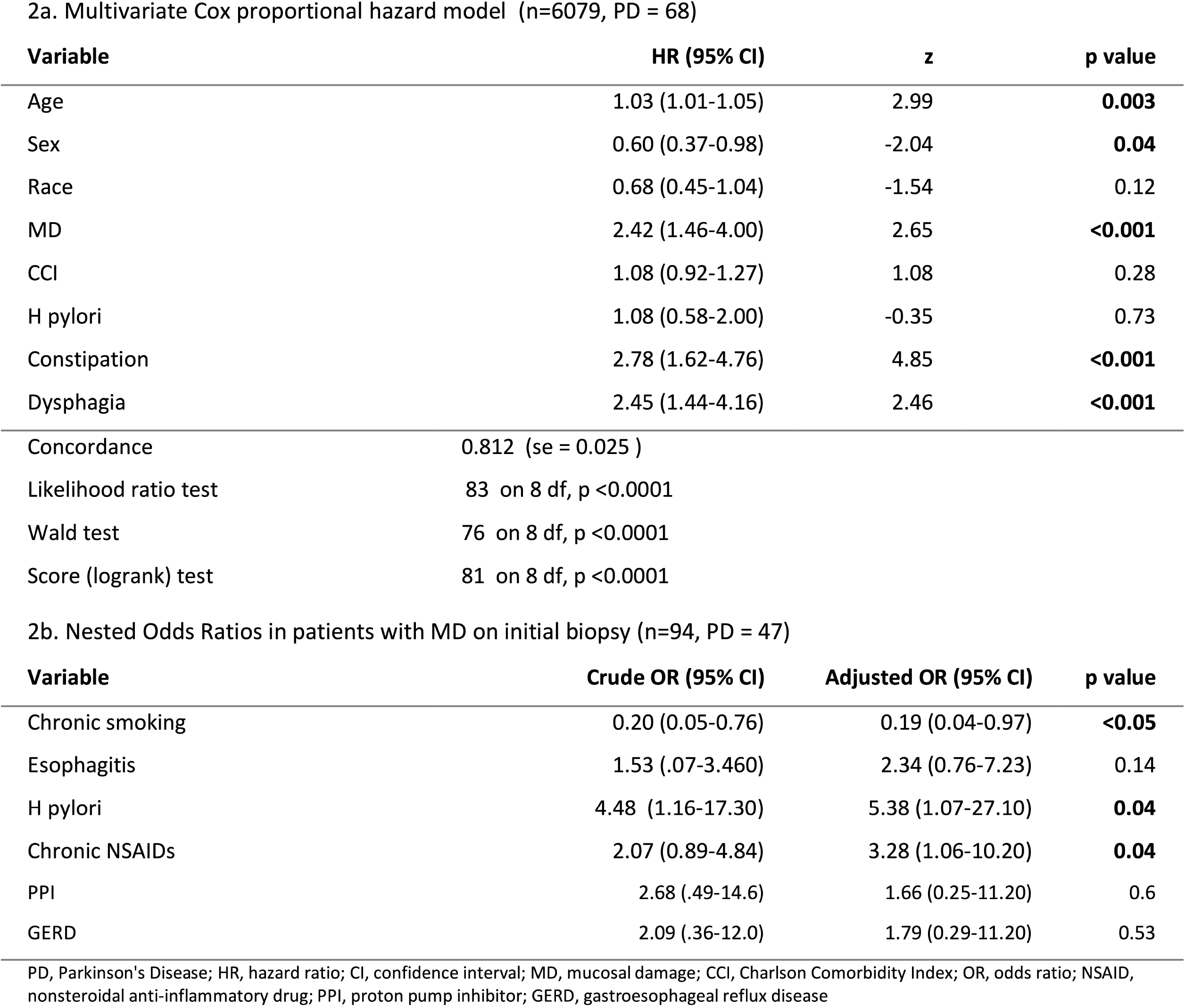
Variables associated with PD Development in patients with upper endoscopy biopsy.

**Figure 3:**
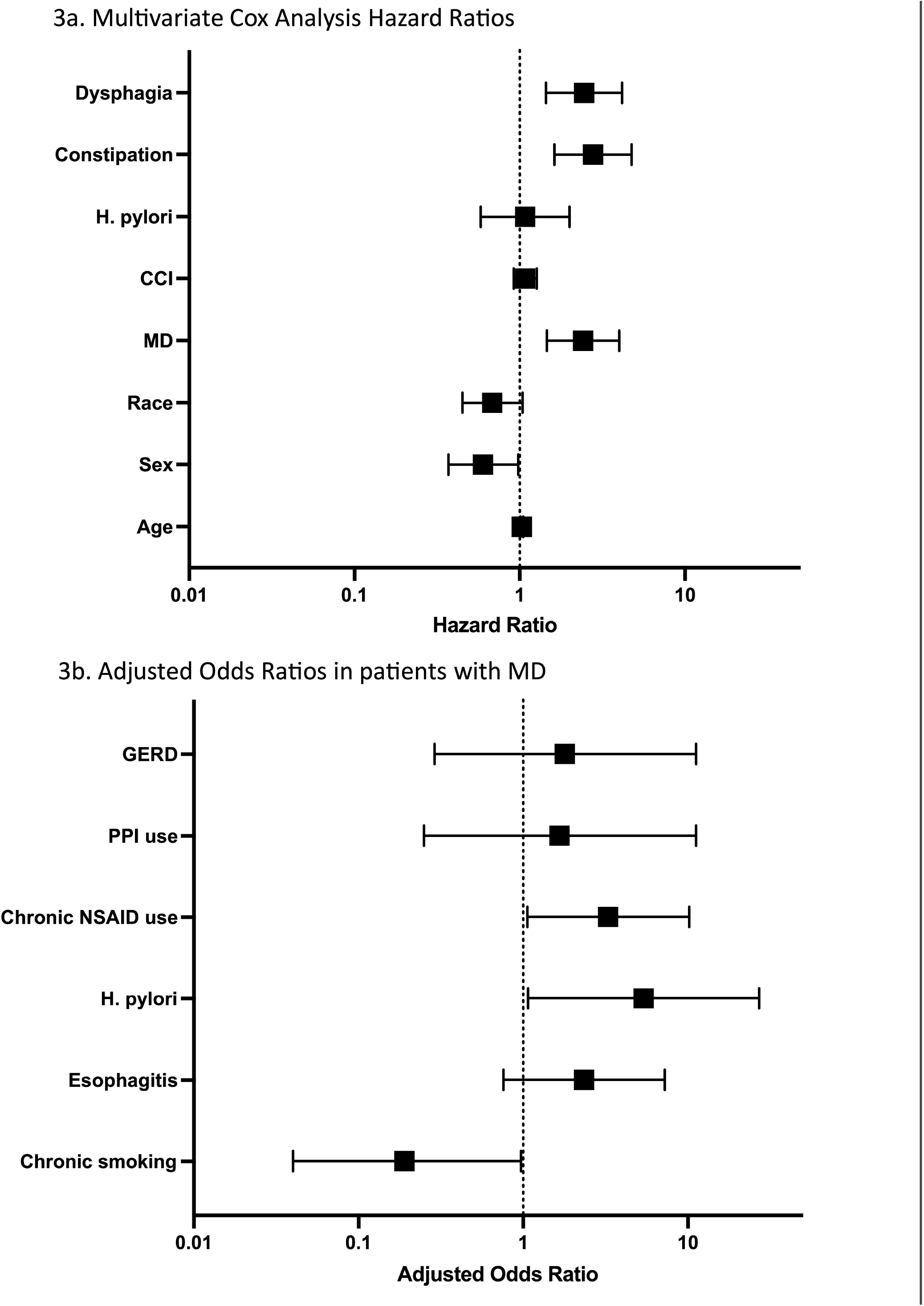
Variables associated with PD in patients with endoscopy Parkinson’s Disease; CCI, Charlson Comorbidity Index; MD, mucosal damage; GERD, gastroesophageal reflux disease; PPI, proton pump inhibitor; NSAID, nonsteroidal anti-inflammatory drug

Within our nested subgroup of patients with MD (**Table 2b, Figure 3b**), presence of *H. pylori* on initial biopsy was linked with a markedly higher probability of developing PD, as indicated by an adjusted odds ratio [aOR] of 5.38 (95% CI 1.07-27.10, p=0.04). Chronic use of NSAIDs also showed a correlation with increased PD risk (aOR 3.28, 95% CI 1.06-10.20, p=0.04).

Conversely, chronic smoking was associated with reduced odds of developing PD in the context of MD (aOR 0.19, 95% CI 0.04-0.97, p<0.05). Esophagitis, PPI use, and GERD were not associated with PD risk.

## Discussion

The findings of our investigation corroborate our hypothesis that GI MD constitutes a significant risk factor for PD onset, reinforcing the theory of a gut-first progression in PD. Our comprehensive analysis indicates a marked escalation in PD risk amongst individuals with MD (IRR 3.00, p<0.0001). This association persists even subsequent to adjustment for established covariates (HR 2.42, p<0.001), underpinning the pivotal role of the gut-brain axis in neurodegenerative conditions and highlighting GI factors as potential early biomarkers and contributors to PD pathogenesis.

An important strength of our study is its access to endoscopic reports and pathologic diagnoses, not merely relying upon ICD-codes for exposure history. Nonetheless, there are some important limitations including our inability to capture diagnoses of PD that were diagnosed and treated outside the MGB system. Given in our large cohort, we were able to identify an incidence of PD comparable to the general population, however, it is likely that we are closely approximating true population burden. Another limitation is that we did rely on ICD-codes for diagnosis of several outcomes, including GERD.

Consistent with prior studies^6^, our investigation did not establish a connection between GERD and PD, though there is well-documented relationship between GERD and MD, and we observed that GERD and PPI use was significantly higher in our MD cohort (Table 1). There are limitations utilizing ICD codes for GERD diagnosis, and likely some insufficiency of what is often now a clinical diagnosis of GERD based on symptoms and empiric response to PPI therapy.^15^ Nonetheless, this lack of a significant correlation between gastroesophageal reflux disease (GERD), proton pump inhibitor (PPI) usage, and PD incidence in our study population challenges the assumption that pH alterations, commonly associated with *H. pylori* infection, are a primary risk factor for PD. This finding necessitates a re-examination of GERD and PPI usage as potential contributors to PD risk.

Our other findings are consistent with existing literature, which identifies age, sex, constipation, and dysphagia as risk factors for PD, thereby demonstrating the reproducibility of our results^3^. Conversely, our study discerns no significant link between esophagitis and PD in patients with MD, suggesting that the relationship between MD and PD risk may be contingent upon either a cumulative effect of various forms of GI inflammation or a synergistic impact of multiple manifestations of MD. This observation intimates that the cumulative burden or the diversity of inflammation sites within the GI tract is instrumental in PD risk elevation, possibly through a threshold effect on the gut-brain axis.

Furthermore, our research unveils a complex interplay between *H. pylori* infection, MD, and the risk of PD. The initial presence of *H. pylori* noted on biopsies correlates with an amplified probability of PD (Adjusted Odds Ratio [AOR] 5.38, p=0.04) but only within the context of MD, indicating a multifaceted interaction where *H. pylori*’s role in PD risk amplification may be contingent on concurrent MD; where the pathogen alone may not be sufficient to elevate PD risk, but its presence in the setting of MD appears to be a significant factor. Indeed, there was no significant difference in *H. pylori* infection rates between MD and non-MDs in our cohort (Table 1). While prevalence of *H. pylori* infection worldwide is approximately 50%, with rates ranging from 35-40% in the US, only about 20% of infected individuals develop related gastroduodenal disorders, including PUD, during their lifetime; individuals with *H. pylori* infection often present asymptomatically^16,17^.

In the context of MD, chronic use of Non-Steroidal Anti-Inflammatory Drugs (NSAIDs) also emerged as a risk factor for PD (aOR 3.28, p=0.04), contributing to the debate over NSAIDs’ role in modulating PD risk. Interestingly, while NSAIDs have been well-established to cause MD through a variety of mechanisms^18^, existing literature is equivocal with regards to the exact role of NSAIDs in modifying PD risk. Some studies cite evidence of reduced PD risk with NSAID use^19^, while other studies, including our findings, indicate no such benefit^11^ and potentially a deleterious impact, particularly in individuals with MD.

Our study also sheds light on the inverse relationship between chronic smoking and PD risk, exclusively within the subset of MD patients (AOR 0.19, p<0.05). While smoking is a well-established risk factor for peptic ulcer disease, consistent with our study, our nested case-control findings supporting a protective role of tobacco use in PD-specific gut pathology This is consistent with prior literature suggesting a dose-dependent inverse correlation between tobacco smoking and PD risk^20-23^. Interestingly, smoking has also been found to lower the risk of ulcerative colitis, possibly due to nicotine’s potential to increase the thickness of the colonic mucosa^24^. Yet this and other protective mechanisms, involving nicotine, genetic polymorphisms, and immunomodulatory signaling, warrant further exploration^20,25-27^, particularly in relation to MD.

In conclusion, our study highlights the necessity for heightened monitoring of patients with MD and the importance of establishing gut biomarkers, given their increased susceptibility to PD. With peptic ulcer disease globally affecting upwards of 8.09 million people^28^, and *H. pylori* infection even more widespread, timely detection and treatment of *H. pylori* infection, along with MD management, may prove crucial to reducing incidence of PD. Further studies investigating the interplay of *H. pylori*, NSAIDs, and smoking, as they relate to different severities of MD, may be crucial for the development of more effective prevention strategies for Parkinson’s Disease.

## Data Availability

All data produced in the present study are available upon reasonable request to the authors

## Acknowledgements

Funded by the American Gastroenterological Association Research Foundation’s Research Scholar Award – AGA2022-13-03

